# Household transmission of SARS-CoV-2 during the Omicron wave in Shanghai, China:a case-ascertained study

**DOI:** 10.1101/2022.09.26.22280362

**Authors:** Zhongqiu Wei, Wenjie Ma, Zhonglin Wang, Jingjing Li, Xiaoming Fu, Hailing Chang, Yue Qiu, He Tian, Yanling Ge, Yanfeng Zhu, Aimei Xia, Qianhui Wu, Gongbao Liu, Xiaowen Zhai, Xiaobo Zhang, Yan Wang, Mei Zeng

## Abstract

**Background:** Since late 2021, the highly transmissible SARS-CoV-2 Omicron variant has driven a new surge of infections across the world. We used a case-ascertained study to determine the features of household transmission of SARS-CoV-2 Omicron variant in Shanghai, China.

**Methods:** We collected detailed information on 323 pediatric cases and their 951 household members in April 2022 during the Omicron outbreak. All household members received consecutively intensive RT-PCR testing for SARS-CoV-2 and routine symptom monitoring within 14 days after exposure to a confirmed case. We described the characteristics of study participants and estimated the transmission parameters. Both secondary infection attack rates (SAR_I_) and secondary clinical attack rates (SAR_C_) among adult household contacts were computed, through which the transmission heterogeneities in infectivity and susceptibility were characterized and the vaccine effectiveness were estimated.

**Results:** We estimated the mean incubation period of SARS-CoV-2 Omicron variant to be 4.6 (median: 4.4, IQR: 3.1-6.0) days and the mean serial interval to be 3.9 (median:4.0, IQR: 1.4-6.5) days. The overall SAR_I_ and SAR_C_ among adult household contacts were 77.11% (95% confidence interval [CI]: 73.58%-80.63%) and 67.03% (63.09%-70.98%). We found higher household susceptibility in females, while infectivity was not significantly different in primary cases by age, sex, vaccination status and clinical severity. The estimated VEs of full vaccination was 14.8% (95% CI: 5.8%-22.9%) against Omicron infection and 21.5% (95% CI: 10.4%-31.2%) against symptomatic disease. The booster vaccination was 18.9% (95% CI: 9.0%-27.7%) and 24.3% (95% CI: 12.3%-34.7%) effective against infection and symptomatic disease, respectively.

**Conclusions:** We found high household transmission during the Omicron wave in Shanghai due to asymptomatic and pre-symptomatic transmission in the context of city-wide lockdown, indicating the importance of early detection and timely isolation of SARS-CoV-2 infections and quarantine of close contacts. Marginal effectiveness of inactivated vaccines against Omicron infection poses great challenge for prevention and control of the SARS-CoV-2 Omicron variant.

## Background

The COVID-19 pandemic caused by SARS-CoV-2 has resulted in unprecedented global health crisis and more than six million deaths worldwide since December 2019 [1]. Despite the increasing natural immunity and vaccine-induced immunity are common in population, the newly emerged Omicron variant, with increased transmissibility and immune escape properties, has rapidly replaced previous strains and driven a new surge of SARS-CoV-2 infections across the world [2, 3]. China maintained local containment through effective border controls and non-pharmaceutical interventions (NPIs) since 2020 and has successfully coped with several importation-linked local outbreaks of SARS-CoV-2 variants [4]. In the meantime, Chinese government spared no efforts to promote countrywide mass COVID-19 vaccination roll-out among adults since April 2021 and among children aged 3-17 years since July 2021 [5,6]. Nevertheless, following the first cluster of Omicron infections detected in late February, 2022, a local epidemic wave caused by Omicron BA.2 sub-lineage hit Shanghai, one of the largest metropolitans with a population of nearly 25 million in China, with widespread community transmission occurred in late March and peaked in April, despite the implementation of mass vaccination and city-wide lockdown [7]. As of May 31, 2022, when the lockdown was lifted, over 0.6 million confirmed cases including 588 deaths were reported in Shanghai [8].

Transmission dynamics of SARS-CoV-2 may potentially evolve over time and vary by settings and with intervention measures. Households are important transmission venues for SARS-CoV-2 [9-11]. A full understanding of the household transmission patterns of SARS-CoV-2 Omicron variant is crucial to plan and adjust the public health responses and target intervention in face of the current challenge of Omicron epidemics. Recently, a few studies from Danish, Norway and the US have reported higher household secondary attack rates (25.1%~52.7%) for Omicron variant than for Delta variant [12-16]. However, accurately determining the household transmission dynamics regardless of symptoms remains challenging, as most studies were based on the analysis of symptom-based screening data, with asymptomatic infections and mild non-medically consulted infections underreported. This challenge can be addressed by studies of close contacts with routine SARS-CoV-2 testing regardless of symptoms to detect asymptomatic and mildly symptomatic cases. As household contacts of SARS-CoV-2-positive cases are likely to be highly exposed to the case and are known to be at high-risk of infection, they are an ideal group shedding lights on SARS-CoV-2 transmission dynamics [17].

Here, we conducted a case-ascertained study to determine the features of household transmission of SARS-CoV-2 Omicron variant in Shanghai, China. In particular, we estimated the distribution of key time-to-event intervals, quantified the household transmission risk and explored the transmission heterogeneities in infectivity and susceptibility. In the meantime, we also assessed the vaccine effectiveness of inactivated COVID-19 vaccines against Omicron infection and symptomatic disease.

## Methods

### Cases and household contacts

A confirmed case is defined as a person with PCR-confirmed SARS-CoV-2 infection, irrespective of clinical signs and symptoms, and further classified as asymptomatic, mild, moderate (non-severe pneumonia), severe and critical case based on both national and World Health Organization (WHO) guidance [18,19]. Pneumonia was diagnosed based on either radiological evidence or typical clinical signs (fever and or cough accompanying with one of the following signs: moist rales, difficulty in breathing, fast breathing, chest indrawing).

A household is defined as two or more people living in the same residence. A household contact is defined as any person who has resided in the same household with a confirmed case for the period from 2 days before to 14 days after the date of symptom onset or laboratory confirmation. As a part of public health monitoring, consecutively intensive RT-PCR testing and routine symptom monitoring within 14 days after exposure to a confirmed case are required for all household contacts every day or every other day [20].

The primary case(s) of a household was defined as confirmed case(s) with a history of community exposure (i.e., exposed to SARS-CoV-2 contaminated environment or contact with a confirmed case in the community). For a household without determined source of infection, we defined the primary case(s) as the first individual(s) who was tested positive with RT-PCR or developed symptoms. Other household members with positive RT-PCR results were defined as secondary cases.

### Study design and participants

This case-ascertained study was conducted in April 2022 at the Children’s Hospital of Fudan University, a designated hospital for management of pediatric COVID-19 cases in Shanghai. During the study period when a large number of pediatric cases of SARS-CoV-2 infection were detected through mass screening, the majority of pediatric cases younger than 5 years old with febrile symptom and all pediatric cases with suspected pneumonia or comorbidities requiring special medical attention were referred to the designated children’s hospitals, while most asymptomatic and milder pediatric cases were transferred to designated isolation facilities for medical observation. We enrolled a total of 335 pediatric cases infected with SARS-CoV-2 Omicron variant who were referred to the Children’s Hospital of Fudan University during April 4 to April 27(Fig. 1). All cases were PCR-confirmed before hospitalization. The confirmed cases are discharged from isolation if the cycle threshold (Ct) value ≥35 for the viral nucleic acid on RT-PCR test [18].

**Fig. 1.**
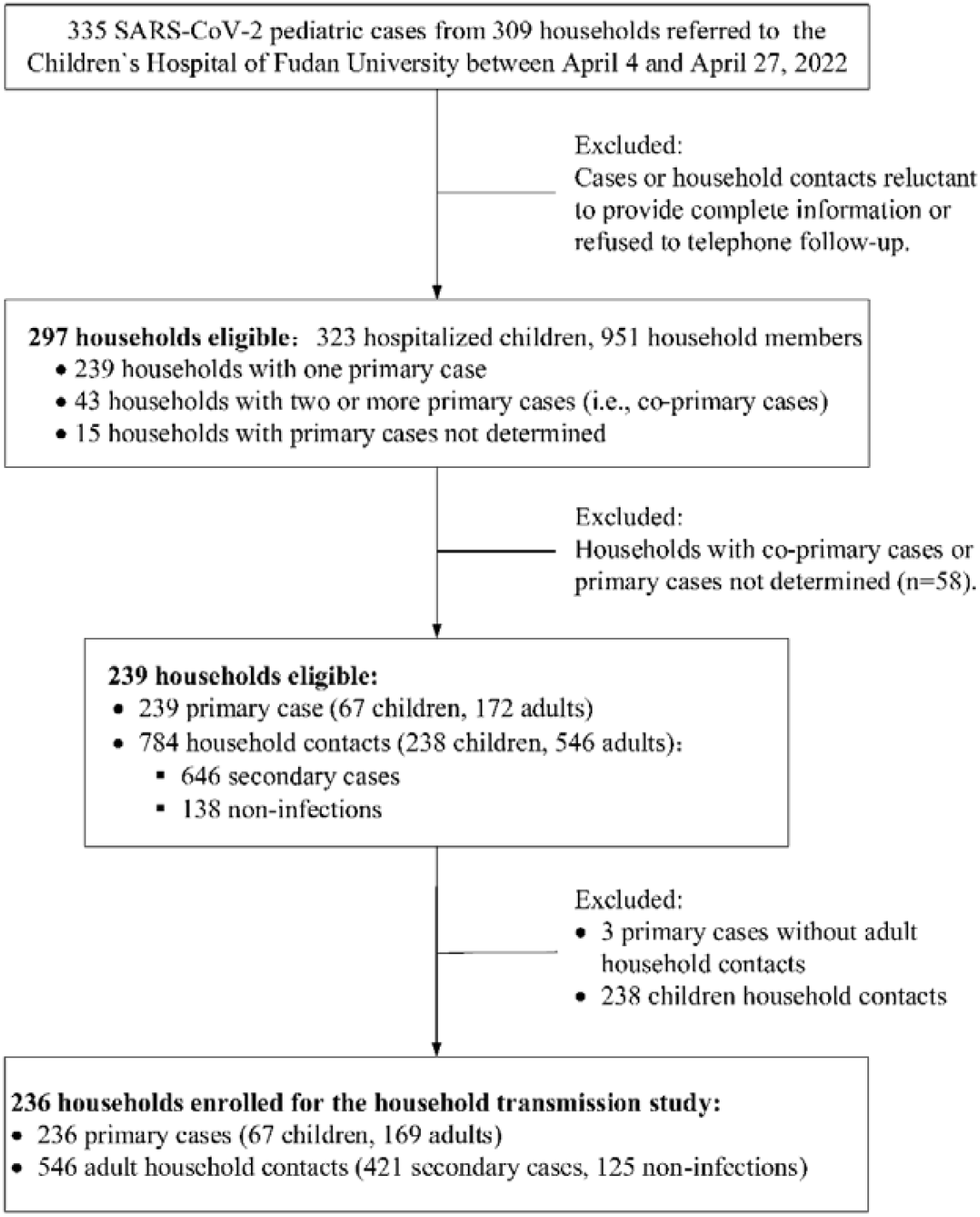
Flow chart describing the procedure for screening study participants.

We further enrolled all household members of the hospitalized children and followed up household cases until one week after discharge. The households were excluded from the study if any of the household members were reluctant to provide the complete information or refused to telephone follow-up. (Fig. 1).

### Data collection

All hospitalized children had at least one household member living together. The household information of each hospitalized case was collected according to case management requirement, including demographics, exposures, vaccination status and symptoms. Clinical data of hospitalized children was extracted from the electronic medical records. Other detailed data on hospitalized children and their household contacts were obtained through face-to-face interview to the accompanying parents using a standard questionnaire (Table S1) during the hospital stay and through the routine telephone follow-up one week after discharge.

### Statistical analysis

We estimated the incubation period (i.e., the period of time from an exposure resulting in SARS-CoV-2 infection to symptom onset) by analyzing cases with clear exposure history. When multiple or sustained exposure was reported, the time interval between the first and last recorded dates of exposure was considered to account for the uncertainty of infection time. We also estimated the serial interval (i.e., the time interval between the onset of symptoms in a primary case and his/her secondary cases). For a secondary case contacts with multiple infections, we randomly selected one as his/her primary case and simulated 100 times to account for potential uncertainties. We fitted three parametric distributions (Weibull, gamma and lognormal) to time-to-event data and selected the best fit based on the minimum Akaike information criterion. The distributions of serial interval were fitted with a shift parameter allowing negative values.

For the household transmission study, we excluded the households with two or more primary cases and the households without determined primary cases from the analysis to avoid potential bias (Fig. 1), as it is possible that a secondary case may be misclassified as a co-primary case. The secondary infection attack rate (SAR_I_) was defined as the number of PCR-confirmed cases detected regardless of symptom among all household contacts of the primary case [17]. The secondary clinical attack rate (SAR_C_) was defined as the number of symptomatic cases detected among all household contacts of the primary case [17]. In this study, there was a potential bias in the estimates of SAR_I_ and SAR_C_ among children household contacts due to the study design that the enrolled households were selected from the families of the confirmed hospitalized pediatric cases. Therefore, we estimated the SAR_I_ and SAR_C_ among adult household contacts to assess the heterogeneities in infectivity and susceptibility. Specifically, the heterogeneities in susceptibility were estimated by the characteristics (e.g., sex and vaccination status) of adult household contacts. The heterogeneities in infectivity were measured by the characteristics (e.g., age, sex, symptom profile and vaccination status) of primary cases (including children and adults).

Comparison between groups was performed using chi-square test. A difference with P <0.05 at two-side was considered to be statistically significant. We estimated the vaccine effectiveness against Omicron infection (VE_I_) and against clinical symptoms (VE_C_) based on the estimates of SAR_I_ and SAR_C_ among adult household contacts with different vaccination status. Specifically, the estimates of VE_I_ were obtained from *VE*_*l,v*_ = 1 − (*SAR*_*l,v*_*/SAR*_*l,u*_),where *v* = 1,2,3, donates the partially, fully and booster vaccinated groups among the adult household contacts, respectively. *SAR*_*l,v*_ donates the secondary infection rate of each vaccinated group and *SAR*_*l,u*_ donates that of the unvaccinated group. Similarly, the estimates of VE_C_ were obtained from *VE*_*C,v*_ =1 - (*SAR*_*C,v*_ */SAR*_*C,u*_), where *SAR*_*C,v*_ donates the secondary clinical attack rate of each vaccinated group and *SAR*_*C,u*_ denotes that of the unvaccinated group. Statistical analysis was preformed using the R software, version 4.0.2; the data were stored and maintained using Microsoft Office Excel 2019.

## Results

A total of 1274 participants from 297 households, including 323 hospitalized children and 951 household contacts, were finally recruited to the study (Fig. 1). Their epidemiological and clinical characteristics were described in Table S2 and Fig. S1. All household cases in this study were non-severe or asymptomatic, except a 7-year-old child, who was critically ill.

We analyzed the period of time from exposure resulting in SARS-CoV-2 infection to disease onset for the 52 symptomatic cases with clear exposure history. We estimated a mean incubation period of 4.6 (median: 4.4, IQR: 3.1-6.0) days, with a standard deviation (sd) of 2.1 days and the 95th percentile of the distribution at 8.3 days (Fig. 2A). The incubation period was well approximated by a Weibull distribution (Table S3). We estimated the time of symptom onset between the 234 transmission pairs. The serial interval followed a best fitted Weibull distribution with an estimated mean of 3.9 (median:4.0, IQR 1.4-6.5) days and a standard deviation of 3.7 days (Fig. 2B, Table S4).

For household transmission study, we excluded 43 households with two or more primary cases, 15 households without primary cases determined and 3 households without adult household contacts (see Method section for details). We finally included 236 primary cases and their 546 adult household contacts for analysis (Fig. 1). The characteristics of the 236 primary cases and their 546 adult household contacts were described in Table 1. Among the 236 household primary cases, 169 (71.61%) were adults and 134 (56.78%) were females. Only 89 (37.71%) of the primary cases reported a clear history of community SARS-CoV-2 exposure, indicating most of the households without a determined source of infection. We found 37.71% of the primary cases were unvaccinated, 2.97%, 35.59% and 23.73% of the primary cases received partial, full and booster vaccination, respectively. Most primary cases (89.83%) were symptomatic. Among the 546 adult household contacts, 421 secondary cases were identified and 366 (86.94%) developed symptoms. The overall SAR_I_ and SAR_C_ among adult household contacts were 77.11% (95% CI: 73.58%-80.63%) and 67.03% (95% CI: 63.09%-70.98%), respectively. We found the infectivity was not significantly different in primary cases with different age, sex, vaccination status and symptom profile (P>0.05, Table 2). For the transmission heterogeneities in susceptibility, we found a higher proportion of females (59.86% vs 38.4%) and a lower proportion of vaccinated individuals (76.72% vs 89.9%) in secondary cases than in uninfected household contacts (Table 1). Accordingly, we found higher susceptibility to SARS-CoV-2 Omicron infection in females (SAR_I_=84%) within household than in males (SAR_I_=68.7%, p<0.001). Similar conclusion was reached when the susceptibility was measured by SAR_C_ (74% for females and 58.54% for males, p<0.001). Unvaccinated adults were associated with the highest risk of household infection (SAR_I_=88.29%) and symptomatic infection (SAR_c_=81.08%), while SAR_I_ could be reduced to 84%, 75.21% and 71.59% (P=0.007), and SAR_C_ could be reduced to 76%, 63.68% and 61.36% (p=0.002), through partial, full and booster vaccination, respectively (Table 3).

**Table 1.** Characteristics of the 236 primary cases and their 546 adult household contacts.

| Characteristics | Primary cases<br>(N=236) | Adult household contacts |  |
| --- | --- | --- | --- |
|  |  | Secondary cases<br>(N=421) | Uninfected contacts<br>(N=125) |
| Age group, years |  |  |  |
| 0-17 | 67 (28.39) | 0 (0) | 0 (0) |
| 18+ | 169 (71.61) | 421 (100) | 125(100) |
| Sex |  |  |  |
| Male | 102 (43.22) | 169 (40.14) | 77 (61.6) |
| Female | 134 (56.78) | 252 (59.86) | 48 (38.4) |
| Community exposure |  |  |  |
| Yes | 89 (37.71) | 0 (0) | 0 (0) |
| No | 0 (0) | 421 (100) | 125(100) |
| Not determined | 147 (62.29) | 0 (0) | 0 (0) |
| Vaccination status* |  |  |  |
| Unvaccinated | 89 (37.71) | 98 (23.28) | 13 (10.4) |
| Partial | 7 (2.97) | 21 (4.99) | 4 (3.2) |
| Full | 84 (35.59) | 176 (41.81) | 58 (46.4) |
| Booster | 56 (23.73) | 126 (29.93) | 50 (40) |
| Symptom status |  |  |  |
| Symptomatic | 212 (89.83) | 366 (86.94) | - |
| Asymptomatic | 24 (10.17) | 55 (13.06) | - |

<sup>\*</sup>Partial vaccination was defined as an individual receiving only one-dose inactivated vaccine. Full vaccination was defined as an individual receiving two doses of inactivated SARS-CoV-2 vaccines for at least 2 weeks. Booster vaccination was defined as a fully vaccinated individual receiving an additional dose of inactivated vaccine for at least 14 days.

**Table 2.** **Infectivity of primary cases**, measured by secondary infection attack rate (SAR_I_) and secondary clinical attack rate (SAR_C_), based on the analysis of 236 primary cases and their 546 adult household contacts^*^.

| Characteristics of primary cases | No. of primary cases<br>( N=236 ) | No. of adult household contacts<br>(N=546) | No. of secondary cases<br>(N=421) | Infectivity measured by SAR<sub>I</sub>, % (95% CI) | P value | No. of secondary cases developing symptoms<br>(N=366) | Infectivity measured by SAR<sub>C</sub>, % (95% CI) | P value |
| --- | --- | --- | --- | --- | --- | --- | --- | --- |
| Age group, years |  |  |  |  |  |  |  |  |
| 0-17 | 67 | 175 | 129 | 73.71 (67.19-80.24) | 0.235 | 112 | 64 (56.89-71.11) | 0.348 |
| 18+ | 169 | 371 | 292 | 78.71 (74.54-82.87) |  | 254 | 68.46 (63.74-73.19) |  |
| Sex |  |  |  |  |  |  |  |  |
| Male | 102 | 240 | 193 | 80.42 (75.4-85.44) | 0.127 | 168 | 70 (64.2-75.8) | 0.225 |
| Female | 134 | 306 | 228 | 74.51 (69.63-79.39) |  | 198 | 64.71 (59.35-70.06) |  |
| Vaccination status |  |  |  |  |  |  |  |  |
| Unvaccinated | 89 | 221 | 162 | 73.3 (67.47-79.14) | 0.066 | 140 | 63.35 (57-69.7) | 0.240 |
| Partial<sup>#</sup> | 7 | 12 | 7 | - |  | 6 | - |  |
| Full | 84 | 187 | 146 | 78.07 (72.14-84) |  | 131 | 70.05 (63.49-76.62) |  |
| Booster | 56 | 126 | 106 | 84.13 (77.75-90.51) |  | 89 | 70.63 (62.68-78.59) |  |
| Clinical severity |  |  |  |  |  |  |  |  |
| Symptomatic | 212 | 480 | 373 | 77.71 (73.98-81.43) | 0.455 | 329 | 68.54 (64.39-72.7) | 0.060 |
| Asymptomatic | 24 | 66 | 48 | 72.73 (61.98-83.47) |  | 37 | 56.06 (44.09-68.03) |  |

\* Households with co-primary cases or primary cases not determined were excluded from this analysis. We assessed the infectivity of primary cases among their adult household contacts to avoid potential bias due to the study design (detailed in Method Section). <sup>#</sup> The SAR<sub>I</sub> and SAR<sub>C</sub> among adult household contacts of a partially vaccinated primary case were not estimated due to the extremely small sample size (i.e., 12 household contacts corresponding to 7 primary cases)

**Table 3.** **Susceptibility of adult household contacts**, measured by secondary infection attack rate (SAR_I_) and secondary clinical attack rate (SAR_C_), based on the analysis of 546 adult household contacts from 236 households^*^.

| Characteristics of adult household contacts | No. of adult household contacts | No. of secondary cases | Susceptibility measured by SAR<sub>I</sub>, % (95% CI) | P value | No. of secondary cases developing symptoms | Susceptibility measured by SAR<sub>C</sub>, % (95% CI) | P value |
| --- | --- | --- | --- | --- | --- | --- | --- |
| Overall | 546 | 421 | 77.11 (73.58-80.63) | - | 366 | 67.03 (63.09-70.98) | - |
| **Sex** |  |  |  |  |  |  |  |
| Male | 246 | 169 | 68.7 (62.9-74.49) | <0.001 | 144 | 58.54 (52.38-64.69) | <0.001 |
| Female | 300 | 252 | 84 (79.85-88.15) |  | 222 | 74 (69.04-78.96) |  |
| **Vaccination status** |  |  |  |  |  |  |  |
| Unvaccinated | 111 | 98 | 88.29 (82.31-94.27) | 0.007 | 90 | 81.08 (73.79-88.37) | 0.002 |
| Partial | 25 | 21 | 84 (69.63-98.37) |  | 19 | 76 (59.26-92.74) |  |
| Full | 234 | 176 | 75.21 (69.68-80.75) |  | 149 | 63.68 (57.51-69.84) |  |
| Booster | 176 | 126 | 71.59 (64.93-78.25) |  | 108 | 61.36 (54.17-68.56) |  |

\* Households with co-primary cases or primary cases not determined were excluded from this analysis. We assessed the susceptibility among adult household contacts to avoid potential bias due to the study design (detailed in Method Section).

Full vaccination was 14.8% (95% CI: 5.8%-22.9%) and 21.5% (95% CI: 10.4%-31.2%) effective against Omicron infection and symptomatic disease. The estimated VE of booster vaccination was 18.9% (95% CI: 9.0%-27.7%) against Omicron infection and 24.3% (95% CI: 12.3%-34.7%) against symptomatic disease. By contrast, partial vaccination has no significant effect on preventing Omicron infection (4.9%, 95%CI: −14.4%-20.8%) and symptomatic disease (6.3%, 95%CI: −18.9%-26.1%) (Table 4).

**Table 4.** Effectiveness of inactivated vaccines (VE) against SARS-CoV-2 Omicron infection and symptomatic disease.

| Vaccination status | VE against infection | VE against symptomatic infection |
| --- | --- | --- |
| Unvaccinated | Ref | Ref |
| Partial | 4.9 (-14.4, 20.8) | 6.3 (-18.9, 26.1) |
| Full | 14.8 (5.8, 22.9) | 21.5 (10.4, 31.2) |
| Booster | 18.9 (9.0, 27.7) | 24.3 (12.3, 34.7) |

## Discussion

This study of household transmission patterns is based on a well-designed case-ascertained study during the Omicron wave in Shanghai, China, with detailed household investigations and consecutively intensive RT-PCR testing. Our results showed high risk of household transmission due to the transmission from pre-symptomatic and asymptomatic infections, despite the implementation of city-wide lockdown and centralized isolation/quarantine of cases and close contacts in hospitals or designated facilities. We observed no significant difference in transmissibility between symptomatic and asymptomatic individuals and between age, sex and vaccination groups, while susceptibility to Omicron infection among female household contacts was higher than males. Our findings also implied marginal effectiveness of inactivated vaccines against Omicron infection and symptomatic diseases, although inactivated vaccines may show high effectiveness against severe outcomes [18-20].

In this study, detailed information on exposures and symptoms of the study participants was collected through in-depth household investigations, allowing us to provide robust estimation of several key time-to-event distributions. We observed a mean incubation period of 4.6±2.1 days for Omicron variant, slightly longer than prior estimates for Omicron (3.0-3.6 days) [21-25] while shorter than that of the ancestral strain (6.3 days)[26]. The 95th percentile of the incubation period distribution was at 8.3 days, suggesting the feasibility of a shorter quarantine period for close contacts or population at risk. Additionally, studies from Spanish, Netherlands, South Korea, Belgium and the US showed shorter serial intervals for Omicron, with the mean estimates ranging from 2.75-4.8 days [12,25,27,28,29,30,31]. In agreement with prior findings, we observed a mean serial interval of 3.9±3.7 days, falling within this interval. Shortened serial interval suggested increased transmissibility and growth advantage of Omicron variant, making timely contact tracing more challenging [32].

Omicron infection resulted in high attack rates among household contacts in this investigation. We estimated the overall SAR_I_ among adult household contacts to be 77.11%, around 2.5-6 times higher than previous estimates (13.2%~31.6%) in Wuhan, Zhejiang, Shenzhen, Guangzhou and Beijing during the first COVID-19 wave in China when the national lockdown was implemented [10, 33-36], consistent with prior studies indicating increased transmissibility of Omicron to preexisting variants [13,15, 37]. The overall estimates of SAR_C_ among adult household contacts in our study were 67.03%, higher than that reported in the US (52.7%), Danish (31%) and Norway (25.1%) [15-17]. This may be partially explained by the longer duration and higher frequency of contacts between household members during the lockdown period, as well as the circulation of more transmissible Omicron BA.2 sublineage in Shanghai [16]. More particularly, our investigation captured all potential household secondary infections as centralized isolation was required for all household contacts for medical observation and intensive RT-PCR testing regardless of symptom. Besides, despite strict NPIs were implemented in Shanghai (e.g., city-wide lockdown, stay-at-home order, mass testing and isolation/quarantine of all SARS-CoV-2 infections and close contacts), our study showed that silent transmission from pre-symptomatic and asymptomatic infections largely reduced the impact of interventions on stopping the household transmission, stressing the importance of early detection and timely isolation of the confirmed cases and quarantine of their contacts.

During the Omicron wave, substantial increase in pediatric cases of COVID-19 was reported in the US [38]. However, the role of children in Omicron transmission has yet to be fully understood. We observed similar high infectivity in pediatric cases (aged 0-17 years) and in adults (aged 18+ years), indicating that children played an equal role in Omicron transmission in household as adults. We found females were more susceptible to Omicron infection in household than males, in line with the finding reported in an early study from Wuhan [10]. Part explanation was that females are more likely to be the caregivers for the sick cases in households. Our finding also demonstrated the similar high-level transmission rate from symptomatic and asymptomatic primary cases, which implies that symptom-based surveillance is insufficient to prevent and control of COVID-19 epidemic, posing great challenge for prevention and control of Omicron transmission. Of particular note, we observed significantly higher susceptibility to Omicron infection for unvaccinated household contacts, consistent with the findings reported in the latest studies from the US, Danish and Norway [37,12-15]. The estimated VEs against Omicron infection and symptomatic disease was 14.8% and 21.5% for fully vaccination, and 18.9% and 24.3% for booster vaccination. An update meta-analysis based on 4 household transmission studies from Danish, Norway and the US reported that the effectiveness of mRNA vaccines for fully vaccinated contacts was 18.1% [37], which is similar to our findings. The marginal VEs against Omicron infection and mild disease suggest significant immune escape of Omicron variant to vaccine-induced antibody protection and waning vaccine immunity over time [39, 40]. However, the role of the current COVID-19 vaccines remains valuable in minimizing the direct disease burden of SARS-CoV-2 Omicron variant because VE estimates against the Omicron variant remain higher for severe disease in the majority of studies [40]. For severe disease caused by Omicron variant, VE of the primary series showed little decline over six months and the first booster dose vaccination improved VE (≥70%) following three to six months from a booster dose [40].

Household transmission patterns are somewhat heterogeneous across studies. The accuracy of the results may be affected by a high degree of methodologic heterogeneity with respect to method and frequency of testing for diagnosis of contacts, isolation of cases and duration of follow-up. A major strength of this study is that we captured all secondary symptomatic and asymptomatic infections of the recruited households as all household members received consecutive RT-PCR testing for SARS-CoV-2 after a primary household case was identified. The estimation of VE is more objective because exposure risk and contact pattern of household individuals are equal and homogeneous relative to the population-based observational study. However, our study is not without limitations. First, despite in-depth household investigation and follow-up of each case, we could not always reconstruct the entire transmission chain and fully avoid recall bias in individual records. We tried to collect information on source of exposures for each household to avoid potential bias, but there are still some households without determined source of infection. The primary cases of these households were defined as the first household members with positive RT-PCR testing results or the sign of COVID-19 symptoms, which may misclassify the primary and secondary cases of a households. Second, due to the study design (i.e., the study households were selected from those of the confirmed pediatric cases), we can only estimate the transmission risk among adult household contacts, the susceptibility of household children cannot be assessed. Third, we did not collect specific age of household contacts, only classified them as children (i.e., 0-17 years) and adults (i.e., 18+ years). Further investigations with detailed age information could help provide age-specific infectivity and susceptibility of Omicron variant. Multivariate analysis adjusted for age, sex and other related factor should be conducted to estimate the vaccine effectiveness.

## Conclusions

In conclusion, high household transmission during the Omicron wave in Shanghai indicates the importance of early detection and timely isolation of SARS-CoV-2 infections. Marginal effectiveness of inactivated vaccines against Omicron infection poses great challenge for prevention and control of the SARS-CoV-2 Omicron variant, implying the necessity of optimizing vaccine strategies.

## Data Availability

All data produced in the present study are contained in the manuscript and its supplementary information files

## Abbreviations

SAR_I_: Secondary infection attack rate
SAR_C_: Secondary clinical attack rate
CI: Confidence interval
NPIs: Nonpharmaceutical interventions
WHO: World Health Organization
Ct: Cycle threshold
VE: Vaccine effectiveness
VE_I_: VE against Omicron infection
VE_C_: VE against clinical symptoms
sd: standard deviation

## Acknowledgments

We thank Professor Hongjie Yu for his critical comments on the manuscript. We also thank all participating children, parents and grandparents for their collaboration with investigation.

## Ethics approval and consent to participate

Data collection and analysis during the field investigation was a part of public health outbreak investigation. This study was approved by the hospital institutional ethical review board and participant’s informed consent was waived.

## Authors’ contributions

MZ conceived, design and supervised this study. YW designed this study and did statistical analyses. ZW, ZW, JL, XF, HC, YQ, HT, YG, YZ, and AX collected the data and followed up children and their household members. WM and JL pooled the data. YW, WM and ZW did statistical analyses under the supervision of MZ. MZ, YW, ZW, ZW and WM wrote the manuscript, made tables and figures. ZW, ZW and WM edited the submitted manuscript. GL, XZ and XZ provide advice on management of patients and implementation of the study. All authors approved the manuscript for publication.

## Declarations

### Availability of data and materials

All data generated or analyzed during this study are included in this published article and its supplementary information files.

