## Supplementary Materials for "Household transmission of SARS-CoV-2 during the Omicron wave in Shanghai, China:a case-ascertained study"

for

**List:**

**eTable 1. Questionnaire.**

**eTable 2. Epidemiological and clinical characteristics of the 323 index children and their 951 household members.**

**eFig. 1. Vaccination status of the 323 hospitalized children and their 951 household members.**

**eTable 3. Estimates of the incubation period based on the analysis of 52 cases from 28 households.**

**eTable 4. Estimates of the serial interval based on the analysis of 234 transmission pairs.**

**eTable 1. Questionnaire.**

| **Case initial reporting form - for confirmed cases** | |
| --- | --- |
| **Basic information** |  |
| Case ID |  |
| Case name |  |
| Age (years, months) | years months  ☐ Unknown |
| Sex | ☐ Male ☐ Female |
| Household size |  |
| Telephone (mobile) number |  |
| Date of first interview (dd/mm/yyyy) | / / |
| Date of follow up (dd/mm/yyyy) | / / |
| **Exposure history** |  |
| Exposure place | ☐ Community exposure  ☐ Household exposure  ☐ Other places: |
| Exposure start date (dd/mm/yyyy) | / / |
| Exposure end date (dd/mm/yyyy) | / / |
| **Vaccination status** |  |
| Vaccination history | ☐ Yes ☐ No |
| Manufacture of the vaccines |  |
| Doses of vaccination | ☐ Unvaccinated ☐ Partial ☐ Full ☐ Booster |
| Date of the last vaccination (dd/mm/yyyy) | / /  ☐Unknown |
| **Clinical information** |  |
| Date of first positive result of nucleic acid testing or antigen testing (dd/mm/yyyy) | / / |
| Clinical severity | ☐ Asymptomatic  ☐ Mild  ☐ Moderate  ☐ Severe  ☐ Critical |
| Date of symptom onset (dd/mm/yyyy) | / /  ☐ Asymptomatic infection ☐ Unknown |
| Date of admission (dd/mm/yyyy) | / / |
| Date of discharge (dd/mm/yyyy) | / / |
| **Symptoms:** |  |
| Fever (≥37.5°C) or history of fever | ☐ Yes ☐ No ☐ Unknown  If Yes, please specify fever spike: °C and fever duration: days |
| Cough | ☐ Yes ☐ No ☐ Unknown |
| Nasal obstruction | ☐ Yes ☐ No ☐ Unknown |
| Nausea | ☐ Yes ☐ No ☐ Unknown |
| Vomiting | ☐ Yes ☐ No ☐ Unknown |
| Diarrhoea | ☐ Yes ☐ No ☐ Unknown |
| Loss of smell (anosmia) | ☐ Yes ☐ No ☐ Unknown |
| Loss of taste | ☐ Yes ☐ No ☐ Unknown |
| Muscle aches | ☐ Yes ☐ No ☐ Unknown |
| **Underlying disease** |  |
| pre-existing condition/co-morbidity | ☐ Yes ☐ No ☐ Unknown  If Yes, please specify: |
| **Imagology and laboratory testing** |  |
| X-ray or CT image characteristics |  |
| White blood cell count (*10^9/L) |  |
| Neutrophil count (*10^9/L) |  |
| Lymphocyte count (*10^9/L) |  |
| Platelet count (*10^9/L) |  |
| C-reactive protein (mg/L) |  |
| **Case initial reporting form - for household contacts** | |
| **Basic information** |  |
| Household Contact ID |  |
| Related hospitalized children |  |
| Relationship with the hospitalized children |  |
| Age (years, months) | ☐ 0-17 ☐ 18+ |
| Sex | ☐ Male ☐ Female |
| Telephone (mobile) number |  |
| Date of first interview (dd/mm/yyyy) | / / |
| Date of follow up (dd/mm/yyyy) | / / |
| **Exposure history** |  |
| Exposure place | ☐ Community exposure  ☐ Household exposure  ☐ Other places: |
| Exposure start date (dd/mm/yyyy) | / / |
| Exposure end date (dd/mm/yyyy) | / / |
| **Vaccination status** |  |
| Vaccination history | ☐ Yes ☐ No |
| Manufacture of the vaccines |  |
| Doses of vaccination | ☐ Unvaccinated ☐ Partial ☐ Full ☐ Booster |
| Date of the last vaccination (dd/mm/yyyy) | / /  ☐Unknown |
| **Clinical information** |  |
| PCR confirmed cases? | ☐ Yes ☐ No |
| Date of first positive result of nucleic acid testing or antigen testing (dd/mm/yyyy) | / / |
| Symptomatic cases? | ☐ Yes ☐ No |
| Date of symptom onset (dd/mm/yyyy) | / /  ☐ Asymptomatic infection ☐ Unknown |
| **Symptoms:** |  |
| Fever (≥37.5°C) or history of fever | ☐ Yes ☐ No ☐ Unknown |
| Cough | ☐ Yes ☐ No ☐ Unknown |
| Nasal obstruction | ☐ Yes ☐ No ☐ Unknown |
| Nausea | ☐ Yes ☐ No ☐ Unknown |
| Vomiting | ☐ Yes ☐ No ☐ Unknown |
| Diarrhoea | ☐ Yes ☐ No ☐ Unknown |
| Loss of smell (anosmia) | ☐ Yes ☐ No ☐ Unknown |
| Loss of taste | ☐ Yes ☐ No ☐ Unknown |
| Muscle aches | ☐ Yes ☐ No ☐ Unknown |

**eTable 2. Epidemiological and clinical characteristics of the 323 index children and their 951 household members.**

| **Characteristics** | **Hospitalized children** | | | **Household members** | |
| --- | --- | --- | --- | --- | --- |
|  | **Non-pneumonia cases**  **(N=279)** | **Pneumonia cases**  **(N=44)** | **Overall**  **(N=323)** | **Infected**  **(N=787)** | **Uninfected**  **(N=164)** |
| **Age group, years** | n (%) | n (%) | n (%) | n (%) | n (%) |
| Median (IQR) | 2 (0.83-4.08) | 2 (0.98-3.5) | 2 (0.83-4) | - | - |
| **0-17** | **279 (100)** | **44 (100)** | **323 (100)** | **32 (4.07)** | **15 (9.15)** |
| 0-1 | 79 (28.32) | 11 (25) | 90 (27.86) | - | - |
| 1-2 | 97 (34.77) | 15 (34.09) | 112 (34.67) | - | - |
| 3-5 | 48 (17.2) | 14 (31.82) | 62 (19.2) | - | - |
| 6-12 | 44 (15.77) | 3 (6.82) | 47 (14.55) | - | - |
| 13-17 | 11 (3.94) | 1 (2.27) | 12 (3.72) | - | - |
| **18+** | **0 (0)** | **0 (0)** | **0 (0)** | **755 (95.93)** | **149 (90.85)** |
| **Sex** |  |  |  |  |  |
| Male | 168 (60.22) | 26 (59.09) | 194 (60.06) | 313 (39.77) | 101 (61.59) |
| Female | 111 (39.78) | 18 (40.91) | 129 (39.94) | 474 (60.23) | 63 (38.41) |
| **Community exposure** |  |  |  |  |  |
| Yes | 22 (7.89) | 5 (11.36) | 27 (8.36) | 101 (12.83) | 0 (0) |
| No | 190 (68.1) | 32 (72.73) | 222 (68.73) | 501 (63.66) | 0 (0) |
| Not determined | 67 (24.01) | 7 (15.91) | 74 (22.91) | 185 (23.51) | 164 (100) |
| **Vaccination status^a^** |  |  |  |  |  |
| Unvaccinated | 241 (86.38) | 41 (93.18) | 282 (87.31) | 186 (23.63) | 21 (12.8) |
| Partial | 7 (2.51) | 0 (0) | 7 (2.17) | 35 (4.45) | 8 (4.88) |
| Full | 31 (11.11) | 3 (6.82) | 34 (10.53) | 332 (42.19) | 75 (45.73) |
| Booster | 0 (0) | 0 (0) | 0 (0) | 233 (29.61) | 60 (36.59) |
| **Underlying disease** |  |  |  |  |  |
| None | 266 (95.34) | 39 (88.64) | 305 (94.43) | - | - |
| Febrile seizures | 6 (2.15) | 2 (4.55) | 8 (2.48) | - | - |
| Solid Tumor | 3 (1.08) | 0 (0) | 3 (0.93) | - | - |
| Congenital heart diseases | 2 (0.72) | 0 (0) | 2 (0.62) |  |  |
| Leukemia | 0 (0) | 1 (2.27) | 1 (0.31) | - |  |
| Rett syndrome | 0 (0) | 1 (2.27) | 1 (0.31) |  | - |
| Crohn’s disease | 1 (0.36) | 0 (0) | 1 (0.31) |  |  |
| Allergic rhinitis | 1 (0.36) | 1 (2.27) | 2 (0.62) | - | - |
| **Clinical severity** |  |  |  |  |  |
| **Symptomatic** | **270 (96.77)** | **44 (100)** | **314(97.21)** | **675 (85.77)** | **-** |
| Mild | 270 (96.77) | 0 (0) | 270 (83.59) | - | - |
| Moderate | 0 (0) | 43 (97.73) | 43 (13.31) | - | - |
| Critical | 0 (0) | 1 (2.27) | 1 (0.31) | - | - |
| **Asymptomatic** | **9 (3.23)** | **0 (0)** | **9 (2.79)** | **112 (14.23)** | **-** |
| **Symptoms** |  |  |  |  |  |
| Fever | 263 (94.27) | 43 (97.73) | 306 (94.74) | - | - |
| Fever spike (℃) | 39.3±0.7 | 39.4±1.1 | 39.3±0.8 | - | - |
| Fever duration (days) | 2 (1-3) | 3 (1-3) | 2 (1-3) | - | - |
| Cough | 107 (38.35) | 25 (56.82) | 132 (40.87) | - | - |
| Nausea/vomiting/  diarrhea | 42 (15.05) | 11 (25) | 53 (16.41) | - | - |
| Stuffy nose | 27 (9.68) | 5 (11.36) | 32 (9.91) | - | - |
| Sore throat | 15 (5.38) | 4 (9.09) | 19 (5.88) | - | - |
| Loss of taste/smelling | 3 (1.08) | 1 (2.27) | 4 (1.24) | - | - |

^a^Partial vaccination was defined as an individual receiving only one-dose inactivated vaccine. Full vaccination was defined as an individual receiving two doses of inactivated SARS-CoV-2 vaccines for at least 2 weeks. Booster vaccination was defined as a fully vaccinated individual receiving an additional dose of inactivated vaccine for at least 14 days.

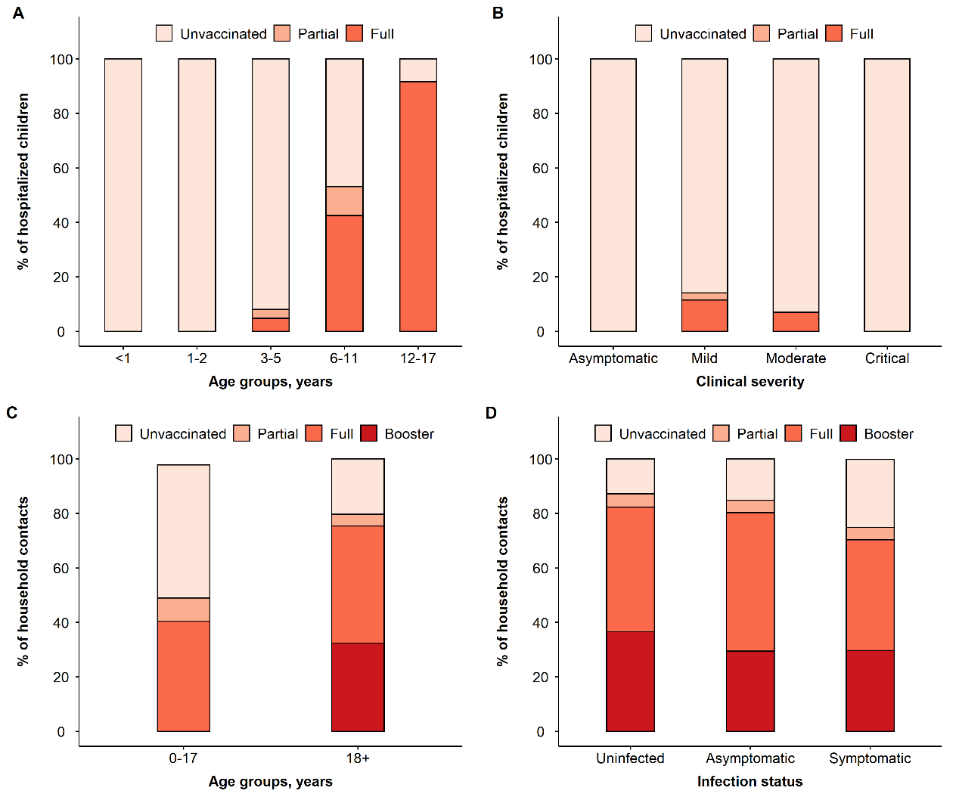

**eFig. 1. Vaccination status of the 323 hospitalized children and their 951 household members. (A).** Vaccination status of the 323 hospitalized children by age. **(B).** Vaccination status of the 323 hospitalized children by clinical severity. **(C).** Vaccination status of the 951 household members by age. **(D).** Vaccination status of the 951 household members by infection status.

**eTable 3. Estimates of the incubation period based on the analysis of 52 cases from 28 households.**

| **Distribution** | **Parameters**  **[mean (SD)]** | **Mean**  **(days)** | **Quantiles**  **(0.025-0.975, days)** | **AIC** |
| --- | --- | --- | --- | --- |
| Gamma | Shape = 3.93 (1.20);  Rate =0.84 (0.29) | 4.67 | 1.25-10.3 | 84.53 |
| Weibull | Shape = 2.36 (0.43)  Scale = 5.19 (0.48) | 4.60 | 1.10-9.03 | 84.14 |
| Lognormal | Meanlog = 1.42 (0.11)  Sdlog = 0.57 (0.10) | 4.87 | 1.36-12.62 | 85.62 |

**eTable 4. Estimates of the serial interval based on the analysis of 234 transmission pairs.**

| **Distribution** | **Parameters^a^**  **[mean (SD)]** | **Mean**  **(days)** | **Quantiles**  **(0.025-0.975, days)** | **AIC** |
| --- | --- | --- | --- | --- |
| Gamma | Shape = 17.01 (0.17);  Rate =1.17 (0.01) | 4.10 | -1.99-11.81 | 106000.4 |
| Weibull | Shape = 4.46 (0.02)  Scale = 15.75 (0.03) | 3.87 | -3.58-10.61 | 104592.4 |
| Lognormal | Meanlog = 2.65 (0.002)  Sdlog = 0.30 (0.002) | 4.34 | -2.68-15.19 | 114545.3 |

^a^ we fitted the distribution with a shift parameter equals to 10.5 days allowing negative serial intervals
